# Identifying patient safety research priorities in a Norwegian hospital setting through a modified James Lind Alliance process

**DOI:** 10.64898/2026.05.04.26352403

**Authors:** Astrid Marie Nysted Berg, Gro Jamtvedt, Dag Karterud, Ida Svege, Sølvi Helseth

## Abstract

**Background:** Patient safety remains a global priority, yet adverse events persist due to gaps in communication, information, training and safety culture. Rapid response systems standardise observation models are widely used to recognise deterioration and guide escalation and response for ward patients in hospitals. A notable gap concerns the role of planning for further care, can improve hospital resource prioritisation as healthcare professionals respond to patients’ deterioration in daily practice. Engaging healthcare professionals as key stakeholders to ensure relevance, we identified unanswered research questions on hospital patient safety and rapid response systems and prioritised the top ten research needs.

**Aim and methods:** We conducted a hospital-tailored, modified James Lind Alliance Priority Setting Partnership (JLA PSP) with healthcare professionals as key stakeholders to identify and prioritise rapid response system related patient safety research needs and evidence uncertainties. The modified JLA process included five stages: (1) establish the Priority Setting Partnership; (2) identify uncertainties; (3) summarise and refine submissions with evidence checks. (4) priority setting; and (5) verify and finalise a top ten list, with evidence checks and project-group oversight throughout.

**Results:** A modified JLA PSP resulted in the stakeholders co-producing a list of research priorities. The top three priorities addressed implementation strategies, intervention effectiveness, and optimising hospital patient safety through clinical protocols and rapid response system activation thresholds. Additional priorities addressed ethical, educational, and organisational factors, highlighting evidence gaps which recognised and responded to patient deterioration and the need for safer transitions across levels of hospital care. The modified JLA PSP was feasible for co-producing a clinically relevant, practice-oriented research agenda.

**Conclusions:** A transparent, systematic, stakeholder-driven process generated hospital patient safety research priorities for rapid response systems that reflect stakeholder needs and target key evidence gaps guiding future research and strengthening patient safety practice in hospitals and, in primary care.

## Background

Patient safety is a key priority in healthcare systems worldwide, with international organisations like the World Health Organization (WHO) promoting initiatives to reduce medical errors and improve patient outcomes [1]. Despite significant advancements, patient safety challenges persist across all levels of healthcare. Adverse events still remain a major cause of death and disability [2–4]. Such events can occur at any stage of care—primary, hospital, or long-term—and often stem from common factors such as communication failures, insufficient information, inadequate training, or poor organisational safety culture [1, 5]. The financial and resource costs of patient harm are substantial, with an estimated 15 precent of hospital activity and spending linked to the preventable adverse events [1]. These incidents represent not only a significant drain on healthcare resources, but also a call for continued improvement in patient safety practices [6, 7].

Healthcare professionals, with their frontline experience, bring critical insights to the challenges and opportunities for improving patient safety in complex hospital settings [8, 9]. Supported by quality improvement organisation, such as the Institute for Healthcare Improvement, and recommended in international guidelines, rapid response systems (RRSs) have been introduced as a key patient safety strategy, but they often vary by institutional policy [8, 10]. Targeted safety initiatives, such as the RRSs have the potential to improve outcomes, reduce harm-related costs, enhance efficiency, and rebuild public trust [7, 11]. The RRSs typically consists of two key components involving healthcare professionals in a hospital setting: the afferent and efferent limbs. The afferent limb focuses on the early detection of patient deterioration, ensuring timely interventions, and activating response assistance when needed, while the efferent limb consists of expert clinicians who provide immediate critical care and facilitate communication to deliver tailored, prompt responses [8, 12]. Rapid response team (RRT) is a key component of a hospital’s rapid response system (RRS) [9, 13]. In hospitals, members of an RRT are expert clinicians—such as specialised physicians and nurses—who are activated to respond to patient deterioration [8, 9, 13]. Across hospitals, RRSs provide round-the-clock cover, ensuring continuous monitoring and rapid response to patient deterioration at any time [8, 14]. RRSs are considered essential for improving outcomes in increasingly complex hospital safety systems [14]. Despite their widespread implementation, challenges remain in optimising their use and evaluating their effectiveness [8]. However, the daily demands on healthcare professionals, combined with the stress of responding to deteriorating patients, may challenge their ability to maintain patient safety [15, 16].

Over the past decade, user involvement in research has grown significantly, highlighting its role in ensuring that research is relevant and useful to health services and thereby shaping practice [17, 18]. Active user involvement—collaborations of patients, healthcare professionals and researchers—enhances research relevance and is associated with improved patient outcomes, including shorter hospital stays, reduced readmissions, better functional recovery and lower mortality rates [9]. However, implementing user involvement in research requires structured frameworks and specific skills; without these, adoption of best practices by healthcare professionals and organisations may be hindered [19]. Engagement methods include advisory and reference groups, workshops, and public events [20]. An important area of user involvement in research is empowering users to identify research needs, especially to prioritise what should be studied, so that study priorities reflect real-world concerns and produce relevant, impactful evidence.

The James Lind Alliance (JLA) is a non-profit initiative established in 2004 [21]. It brings patients, carers and clinicians together in priority setting partnerships (PSPs) to identify and prioritise unanswered healthcare questions and evidence uncertainties, with the result being a top ten priority list of research targets within various fields [21]. By engaging stakeholders meaningfully through a JLA process, these challenges can be addressed effectively, reducing research waste and enhancing the relevance and applicability of healthcare interventions focusing on issues most important to stakeholders [22–24]. This approach integrates two key processes: active user involvement with priority setting partnership (PSP) stakeholders and the identification and validation of evidence gaps through systematic litterature searches [21, 23].

Previous PSPs in primary healthcare has identified unanswered research questions in patient safety [5]. However, to the best of our knowledge, no comparable work has been done in specialist healthcare to identify a top ten list of patient safety research priorities. Engaging healthcare professionals as key stakeholders in research priorities ensures that patient safety challenges, such as those related to RRSs, are addressed with relevant, actionable, and practical solutions in hospital settings. This study seeks to narrow the broad scope of patient safety to focus on identifying unanswered research questions and priorities related to a hospital RRS, ultimately contributing to advancing practices and improving outcomes in a hospital context. This research may provide valuable insights and directions for future patient safety and RRS initiatives and patients’ outcomes.

## Methods

### Aim

The aim of this study was to identify the top ten patient safety research priorities specifically focused on a hospital rapid response system, by actively engaging healthcare professionals as key stakeholders.

### Setting

This study was conducted at one of Norway’s largest emergency university hospitals, which has a capacity of 1000 beds and serves approximately 10 percent of Norway’s population. The study took place over a 9-month period from August 2019 to May 2020 and was approved by the university hospital’s privacy legislation authority (Ref. 2020_124, 20/05868). This hospital provides specialised care in medicine, surgery, mental health, and substance abuse treatment, and is a leading European centre for day surgery, performing approximately 9,000 surgical procedures annually. A key component of the hospital’s patient safety strategy is the implementation of a comprehensive patient safety package of standardised tools and measures for risk assessment. This package specifically focuses on a rapid response system. It includes the National Early Warning Score (NEWS), the proactive course (proACT) training program for preventing and treating life-threatening conditions, and assistance from the rapid response team. NEWS evaluates patient conditions using six vital physiological parameters, namely: respiratory rate, oxygen saturation, temperature, systolic blood pressure, pulse rate, and level of consciousness. These strategies focus on patient deterioration while empowering healthcare professionals within the RRS framework to provide acute medical assistance with the necessary resources. The hospital has a user committee, which serves as a forum for collaboration between former patients, representatives from user organisations, the Cancer Association, the Pensioners Association and the Norwegian Directorate of Health. To include the user committees’ perspectives in this study, the first author organised a pre-study meeting with the Hospital User Committee, which supported the decision to narrow the scope from patient safety work to the hospital RRS.

### A modified James Lind Alliance process

The JLA process is widely recognised for its structured approach to identifying and prioritising research needs, as described in the JLA guidebook [21]. It brings together diverse stakeholders to collaboratively identify and rank uncertainties. Although well established, the JLA process is often time-consuming, conducted on a broad scale, and requires significant resources [25]. In this study, a modified JLA process was developed to suit the specific context of a hospital setting, focus on a defined thematic area (patient safety), and remain manageable within a limited timeframe. The process involved healthcare professionals as key stakeholders and was given a five stages framework: (1) Establish PSP, (2) Identify uncertainties, (3) Summarise, refine and evidence check, (4) Priority setting, (5) Verify and refine into top ten research questions. Fig 1. provides an overview of the JLA PSP process and the modified JLA PSP process. Ethical considerations related to user involvement of the stakeholders were addressed as per institutional guidance. In this study, the REPRISE checklist was used for the priority setting process [26] and the GRIPP2 short form (GRIPP2-SF) reporting checklist was used for documenting user involvement [27]. We applied the Consolidated Criteria for Reporting Qualitative Research (COREQ) checklist in preparing the manuscript [28]. (See supplementary files S2, S3 and S4).

**Fig 1.**
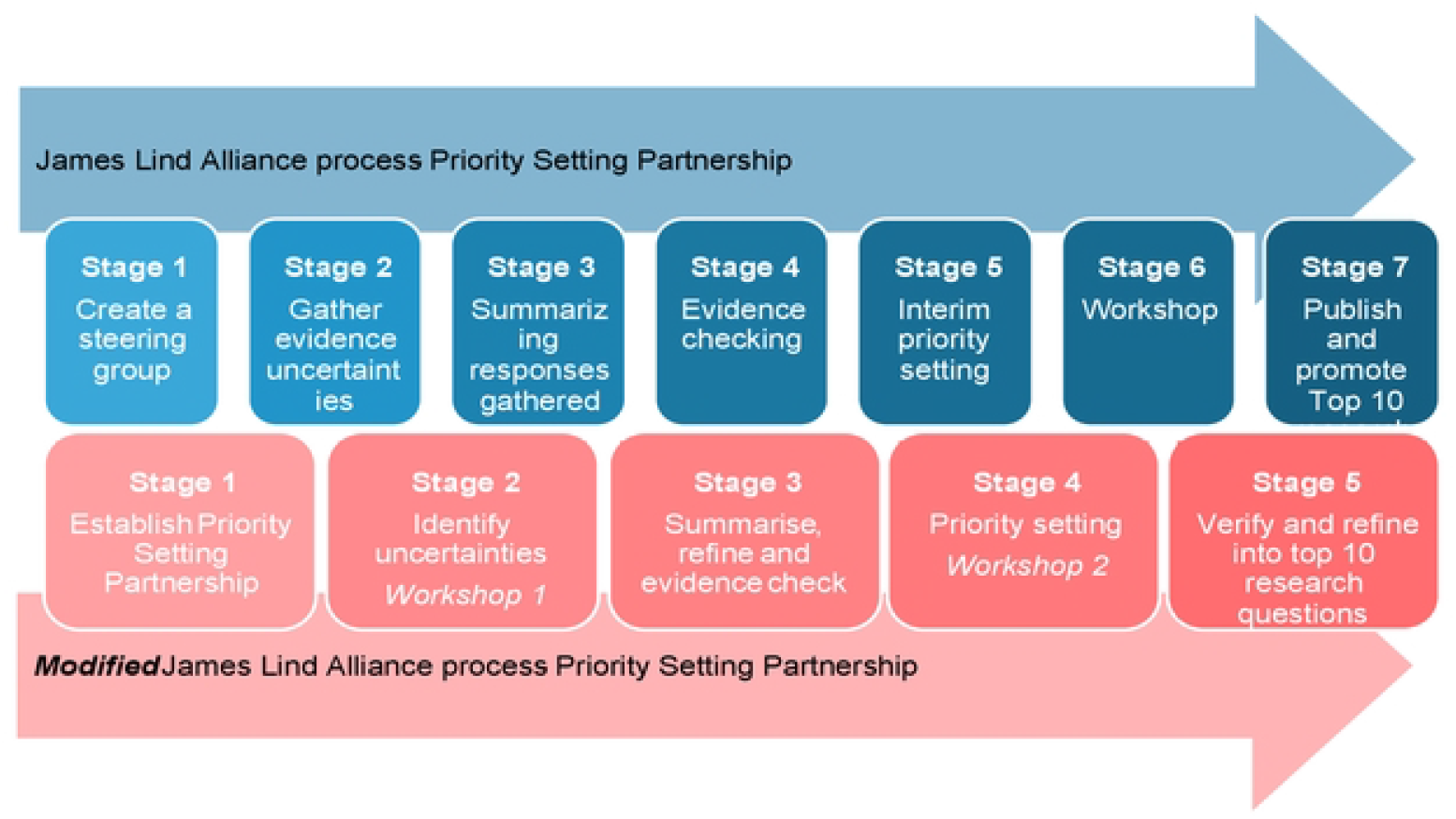
A modified James Lind Alliance process

S1 Fig.1.Fig 1. A modified James Lind Alliance process

The project group of four researchers designed and implemented the modified JLA PSP process by facilitating user involvement, conducting evidence checks of the literature, and supporting decisions at every stage. The following sections will provide a detailed description of the modified JLA PSP process, and the rationale for adaptations made to suit this unique study context.

### Stage 1. Establish the Priority Setting Partnership

The aim of this stage was to identify and recruit key stakeholders for the PSP. The stakeholders consisted of healthcare professionals and researchers organised into a steering group and an expert panel. The steering group played a key role in identifying and prioritising research needs throughout the process. The expert panel provided advisory support through the JLA-process.

A total of 16 participants were recruited by the first author, including 11 for the steering group and 5 for the expert panel (Table 1). Ward leaders from both surgical and medical wards were contacted via email to identify and recruit eligible participants within the scope of the RRS initiative. Inclusion criteria required participants to have at least one year of experience with patient safety initiatives, such as the tools within RRS or being a member of the RRT. To secure eligibility, healthcare professionals without direct involvement in patient safety work, or those unable to commit to the workshops, were excluded. Diversity among key stakeholders was prioritised to capture varied insights as outlined in Table 1. The steering group included nurses and physicians from medical and surgical wards (one of whom was also a section leader), members of the RRT in the Intensive Care Unit, and a former patient from the Hospital User Committee. This former patient expressed interest in participating and, after a pre-study meeting with the user committee, was recruited and included in the PSP. The expert panel consisted of experienced nurses and physicians, with extensive expertise in patient safety, particularly in relation to the hospital’s RRS. It also included senior leaders such as the head of the Surgical Division. Additionally, one stakeholder in the expert panel, an Associate Professor, provided an external perspective as a researcher.

**Table 1.** Stakeholders in the Priority Setting Partnership (PSP)

| PSP stakeholders | Profession/background | Total number |
| --- | --- | --- |
| **Steering group:** | Physicians: 2 Anesthesiologists and members of RRT, 1 neurologist<br>Intensive Care nurses: 1 member of RRT, 1 Special advisor of simulation training<br>Nurses: 1 Gynaecology ward, 1 Gastrology ward, 1 Section leader, Gastrology ward, 1 Medical ward, 1 Orthopaedic ward<br>Former patient, member of Hospital User Committee | 11 |
| **Expert panel:** | Associate Professor, nursing<br>Intensive Care nurse and special advisor of simulation training<br>Senior physician, anaesthesiologist and member of RRT<br>Head of Department from the Surgical Division<br>Section leader from the Intensive Care Unit | 5 |

### Stage 2. Identify uncertainties

The aim of Stage 2 was to identify uncertainties and research needs within the scope of RRS through a workshop with the steering group. To begin with the group adapted the protocol and terms of reference to align with the JLA approach. The workshop then continued with a 10-minute presentation on the JLA-process, followed by 50 minutes of group work and a 30-minute plenary session. Ground rules were established to encourage respect, collaboration and open communication. During the workshop, stakeholders were divided into smaller groups of two and three participants and tasked with identifying and noting uncertainties encountered in daily hospital practice. To guide discussions, they were asked: “What are your questions about patient safety issues related to the general practice, daily use, and management of measures and tools within RRS, the rapid response team, or being a part of the rapid response team?” We dedicated 30-minutes to a plenary session to discuss the workshop, summarise each group’s discussion, and share reflections about the hospital RRS. The workshop, facilitated by the first author with co-researchers assisting through notetaking and observation, resulted in the collection of 87 uncertainties from the steering group.

### Stage 3. Summarise, refine and evidence check

In this stage, the aim was to summarise, refine, and evidence-check the 87 uncertainties identified during Stage 2. The analysis process was inspired by the five stages described in the JLA guidelines: 1. Gather the data, 2. Remove out-of-scope uncertainties, 3. Categorise eligible submissions, 4. Form indicative questions, and 5. Verify the uncertainties [21]. The first author categorised the uncertainties into key themes and analysed them for duplicates. Similar themes were consolidated into indicative questions, while duplicates, comments, and out-of-scope uncertainties were removed between November 2019 and January 2020 as outlined in Table 2. To check the existing evidence, literature searches were conducted in Embase, Cinahl, SweMed +, and Ovid Medline to ensure comprehensive coverage. The search was limited to primary studies and systematic reviews published within the last five years and limited to patient safety and hospital RRS. Following this, uncertainties already answered in the literature were excluded. As a result of Stage 3, the 87 uncertainties were refined into 20 uncertainties for further priority ranking.

**Table 2.** Exclusion categories and number of excluded uncertainties by theme.

| Exclusion category | Excluded uncertainties (n) | Description | Examples of uncertainties |
| --- | --- | --- | --- |
| **Already answered in literature** | 13 | Questions addressed in existing resources e.g., guidelines, systematic reviews or meta-syntheses | “How to use the National Early Warning Score Guidelines in practice” (answered by national guidelines: *National Clinical Guidelines for early detection and rapid response to deteriorating conditions*) |
| **Duplicate** | 28 | Similar uncertainties representing the same purpose or phrasing | “To avoid misunderstandings, how can communication between physicians and nurses improve” and “How can communication between physicians and nurses be improved to avoid miscommunications” |
| **Overlaps** | 22 | Uncertainties overlapping with or addressed by other questions | “What do nurses expect from the rapid response team on the ward” or “What are nurses’ expectations of the rapid response team’s role on the ward” |
| **Out of scope** | 4 | Questions unrelated to the rapid response system | Uncertainties unrelated to the rapid response system |

### Stage 4. Priority setting

The aim of Stage 4 was for the steering group to prioritise the 20 identified uncertainties. This phase allowed the stakeholders to ensure that the research questions aligned with user needs and importance. Each member of the steering group was given the opportunity to individually prioritise up to three questions by marking them on the 20 key uncertainties list. During the workshop, the steering group ranked the questions from “very important” to “unimportant” based on their individual votes and used a consensus process to agree on the top ten list of research priorities. Additionally, the steering group provided input to the project group to ensure the relevance and impact of the final ranked questions. Based on this, the project group finalised a list of the ten most important research priorities related to hospital RRS.

### Stage 5. Verify and refine into the top ten research questions

The aim of Stage 5 called for the expert panel to verify the top ten list of research priorities and refine the identified evidence uncertainties into specific research questions. In this meeting, the expert panel provided advice on methodological approaches for studies addressing these priorities. The panel also identified data and resource limitations in the hospital’s electronic health record system that must be addressed to ensure relevance and robustness of data. Indicative questions were refined using the PICO framework (Population, Intervention, Comparison, Outcome) to ensure clarity and relevance [29, 30]. The project group then finalised the list of the ten most important research priorities as questions (Table 3).

**Table 3.** Top ten research priorities of rapid response systems in a hospital setting.

| Ranking | Top ten research priorities | Steering group voting* |
| --- | --- | --- |
| 1 | How do regular training sessions and case reviews led by the rapid response team impact on the competence, confidence, and collaboration of nurses and physicians in handling deteriorating patients and critical situations? | 10 |
| 2 | How does the involvement of the rapid response team influence resuscitation efforts, ethical decision-making, and the management of end-of-life care for terminally ill patients? | 8 |
| 3 | In what ways do patient safety conditions promote or challenge the implementation of the patient safety package of measures in hospitals? | 5 |
| 4 | Is the current threshold for contacting the rapid response team appropriate, and how do standard protocols and measures impact patient outcomes from hospital admission to discharge? | 3 |
| 5 | How effective is the implementation of the patient safety package of measures in the hospital, and what factors influence its usage and response time by healthcare professionals and wards? | 2 |
| 6 | How can the hospital's patient safety practices be effectively transferred to and implemented within community healthcare services? | 1 |
| 7 | How does interdisciplinary simulation training in proACT courses impact the common understanding, language, resource distribution, and collaboration among nurses, physicians, and the rapid response team in the implementation of the patient safety package of measures? | 1 |
| 8 | What is the optimal composition of the rapid response team (RRT) in terms of specialisation and roles to ensure the best patient outcomes, and should there be multiple teams or specific RRT functions for all intensive care nurses? | 1 |
| 9 | How does the involvement of relatives in the rapid response team (RRT) process impact transparency, trust, and patient outcomes, and what are the best practices for providing information and enabling relatives to request RRT oversight? | 1 |
| 10 | What are the long-term patient outcomes following rapid response team interventions, and how effective are these interventions in improving patient health and recovery? | 1 |

\*Members of the steering group (n=11) ranked top 3 on the key 20 list. The total was 33 votes.

## Results

### The top ten priority list

The stakeholders (PSP) and the project group collaboratively developed the list of research priorities through a ranking process, with the majority of votes concentrated on the top three questions (Table 3). The final list revealed gaps in current research and aimed to inspire future research addressing patient safety issues within the scope of the hospital’s RRS. The list emphasised the need for a well-rounded approach to improve hospital RRS, addressing not only the clinical effectiveness of patient safety interventions but also ethical, educational, and structural aspects of care. Several research priorities focused on implementation, the effects of interventions, and optimising patient safety, including a safety package of measures, clinical protocols, and activation thresholds for hospital RRT. Together, these priorities underscored the need to critically evaluate and improve existing systems to enhance patient outcomes. The diversity of the research priorities reflected engagement from key stakeholders with varied levels of experience, ensuring multiple perspectives. For example, the former patient emphasised clear communication and timely escalation of care; clinicians prioritised reliable patient safety measures, decision-support needs, and workflow support for hospital RRT activation; and managers underscored the need for robust, reliable hospital data and realistic resourcing—collectively guiding investigations into unanswered questions in acute hospital care.

## Discussion

In this study, we identified the top ten research priorities focused on patient safety and the hospital RRS. Hospital healthcare professionals were actively engaged as key stakeholders. The top-ranked research priority questions demonstrate the need to evaluate and explore how simulation, regular training and case reviews led by the RRT can improve the competence, confidence, and collaboration of healthcare professionals in critical situations, reflecting a growing recognition of the importance of professional collaboration in managing complex and critical patient situations effectively. This is consistent with findings from previous research studies showing that systematic team training can be associated with better collaboration and safer patient care pathways [31, 32]. However, these findings need to be further explored.

Our top ten research priorities address universal patient safety challenges related to limited resources, communication barriers, and vulnerable transitions between levels of care, and emphasise operationalising hospital patient safety. In particular, they focus on tangible, RRT-led capability-building and ethical challenges, including regular training and case reviews. These priorities strengthen healthcare professionals’ competence, confidence, and collaboration in recognising and managing deterioration across wards or departments and at the clinician-patient interface, providing clear targets for further research and improvement. Across these areas, tools and strategies to measure, monitor, and improve patient safety remain limited, underscoring the broad applicability of our findings, particularly for diagnostic processes and care transitions [33]. The emphasis on communication, escalation, and data quality is also relevant in primary care, supporting earlier detection, timely escalation, and safer handovers between community and hospital health services.

One priority called for evidence to bridge the gap between hospital and community healthcare services, emphasising the importance of continuity of care and transferable patient safety practices across settings. For comparison, the JLA PSP on patient safety in primary care by Morris et al. [5] highlighted similar themes around communication, transition of care and improving understanding of patient whose safety is vulnerable. Conducted on a broader scale and directly engaging patients, carers, and community stakeholders, their PSP highlighted gaps affecting underserved groups. In contrast, our hospital’s RRS top ten priorities center on operational delivery of safety. PSP stakeholders prioritised evidence on RRT-led training and case review, explicit escalation thresholds and protocols, team composition and response time for patient deterioration, measurements and monitoring, and the transfer of hospital practices to community services. These differences reflect scope and stakeholder composition. Our top ten priorities are based on PSP stakeholder’s experiences from hospital RRS and patient safety work in a Norwegian setting with strong representation of system and service-delivery concerns, whereas Morris et al. [5] engaged a broader primary-care community of patients and carers. The primary care PSP placed greater weight on patient-and community-facing needs, equity for vulnerable populations, and continuity and understanding of safety in an everyday primary care context, elevating priorities related to access, vulnerability, and whole-person care.

It is notable that ethical research questions rank highly in our list of research priorities, underscoring how closely these priorities mirror daily challenges experienced in hospital practice. The second-ranked research priority brings attention to the ethical complexities in decision-making and end-of-life care, indicating an urgent need to investigate how the hospital’s RRT can support healthcare professionals in navigating these morally and emotionally challenging situations, and ensuring patients-centred care. While existing studies provide valuable insights, significant gaps persist in end-of-life-care, especially around cultural competence, standardised educational programs and nurses’ contributions regarding their role in hospital RRS and patient safety improvement [8, 12, 34]. By focusing on priorities identified by frontline hospital healthcare professionals, our work ensures actionable and context-sensitive patient-safety findings, consistent with prior studies showing that co-produced, stakeholder-led priority setting improves relevance and uptake in hospital settings [13, 24, 35, 36].

Our modified JLA PSP process demonstrates how JLA principles can be adapted for smaller-scale projects and applied across patient safety contexts [37, 38]. This pragmatic adaptation addresses a gap in the literature and emphasises user involvement, with stakeholder engagement sustained beyond project completion, demonstrating a robust, collaborative process. Our modified JLA PSP process facilitated broad involvement and collaboration, linking research more closely to clinical practice by engaging stakeholders with diverse roles and hands-on experience in hospital RRS research prioritisation. In doing so, it brought research closer to practice by foregrounding experiential knowledge and ensuring that the priorities reflected real-world needs [18]. User participation in health research parallels the democratisation of the health service and rising expectations for patient and public involvement [38]. To reduce research waste, questions must be relevant to users, grounded in evidence gaps identified from comprehensive evidence syntheses, and not already answered [22]. While PSPs often include multiple participants representing different user groups, including former patients and relatives [5, 35], this study focused on hospital RRS and health professionals with hands-on experience, managing deteriorating patients in hospital settings, and senior leaders, ensuring a broad range of expertise. Despite including a former patient as a stakeholder, the modified JLA PSP may still have deprioritised patients’ top three priorities, as excluding patients and carers may have led to missed evidence gaps particularly relevant to them, for example, communication with families when patients deteriorate. Stakeholder feedback indicated that the modified JLA PSP was feasible and important to both the former patient and the healthcare professionals, who valued the opportunity to shape research priorities directly relevant to their daily practice. For many hospital healthcare professionals, participation in the modified JLA PSP was a novel approach which was well received, and participants were motivated to be involved in similar work in the future.

Using a modified JLA PSP secured data availability and methodological robustness and highlighted the need to strengthen RRS knowledge and experience. Early scoping of RRS enabled an efficient process, guided by a steering group and informed by an expert panel and project team, integrating professional and user perspectives to generate actionable RRS research priorities. We recognise potential trade-offs in identifying research needs through a modified JLA process. Because priorities reflect the context-bound perspectives of those involved, emergent or minority views may be under-represented, and group dynamics can privilege dominant voices. We mitigated these risks through a transparent, five-stage, participant-led, modified JLA PSP process. This modified JLA PSP can be adapted for small-scale contexts to identify actionable patient safety research priorities for RRSs [24, 37, 38].

By focusing on stakeholders’ priorities, we propose a targeted agenda to advance patient safety and strengthen hospital RRS improvements. Achieving this requires collaboration among healthcare professionals, patients, policymakers, and researchers [10, 38, 39]. It also underscores the importance of well-defined roles, adequate resources, and training to meet increasing care demands [12, 13, 39]. This work addresses persistent evidence gaps on how hospital RRS implementation shapes interprofessional practice, as highlighted by Allen et al. [14] and aligns with national calls from health authorities, hospital leaders, and clinicians for systematic evaluations before introducing new hospital technologies [37]. Engaging healthcare professionals with diverse experience revealed evidence gaps, most notably the effect of hospital RRT activation thresholds on patient outcomes, highlighting areas for further exploration [18, 39, 40].

A strength in the hospital prioritisation exercise included the first author’s established networks and field familiarity. This enabled efficient recruitment of user participants to the steering group and expert panel and broad representation of healthcare professionals. The organisational anchoring and the hospital’s sense of ownership enhanced the project’s relevance and integration within the organisation. User representation included both healthcare professionals (experts in their field) and members of the Hospital User Committee (service users who could act as experts), providing a comprehensive perspective. Familiarity with patient safety work ensured that priorities reflected operational needs, workflows and constraints, offering practice-grounded insight that is directly applicable in a hospital setting. Existing relationships expedited coordination across wards and departments. While this could pose a challenge if professionals felt obligated to participate, it also provided the strength of knowing how to effectively engage with them.

However, proximity to the field and existing relationships may have created a sense of obligation to participate, introducing potential bias and a risk of blind spots. Consistent with JLA principles, meaningful user involvement, combining professional expertise with service-user perspectives, linked research prioritisation closely to clinical practice. Including healthcare professionals as user representatives, while valuable, can skew perspectives toward professional interests unless balanced by broader service-user input. Hospital leadership may hold interests that differ from those of the researcher, underscoring the need for critical reflection on influence and decision-making. The first author’s pre-understanding, hypotheses, and professional and theoretical perspectives on patient safety, were advantageous for navigating the setting but may have limited recognition of new insights [41]. Strong organisational anchoring may reduce transferability to contexts with different structures, cultures, or stakeholder compositions.

## Conclusion

Through a transparent and modified JLA PSP process, an experienced group comprising a former patient, health professionals and managers were identified and co-produced a valuable list of research priorities for patient safety and hospital RRS. This modified JLA PSP process facilitated effective engagement of healthcare professionals throughout, ensuring that the priorities were grounded in practice. The resulting list of research priorities creates actionable pathways spanning clinical improvements of hospital RRS, ethics, education and system structure, with PSP stakeholder consensus encompassing the top three priority questions. These priorities provide a practical opportunity to evaluate and optimise hospital RRS and a standardised patient safety package of tools and measures for risk assessment, decision support and RRT activation to improve patient outcomes. We intend these results to guide future research relevant to practice, address key evidence gaps, and strengthen meaningful stakeholder involvement. We anticipate that the top ten list of research priorities may inform and motivate researchers and funders. The first two priorities are already being advanced in a study on patient safety and RRS in a Norwegian hospital.

## List of abbreviations

COREQ: Consolidated Criteria for Reporting Qualitative Research
GRIPP2-SF: Guidance for Reporting Involvement of Patients and the Public–Short Form
JLA: James Lind Alliance
PSP: Priority Setting Partnership
REPRISE: Reporting guideline for Patient and Public Involvement in research
RRSs: Rapid Response Systems
RRS: Rapid Response System
RRT: Rapid Response Team

## Declarations

### Ethics approval and consent to participate

This study adhered to the ethical principles for medical research involving human subjects as outlined in the Declaration of Helsinki [42] and received approval from both the Norwegian Agency of Shared Services in Education and Research (NSD-472886) and the privacy legislation authority at Akershus University Hospital (Ref. 2020_124, 20/05868). The Regional Committee for Medical & Health Research Ethics, Section A, South-East Norway, determined that the research project fell outside the scope of the Act on Medical and Health Research (Ref.172391). Prior to participating in the workshops, all stakeholder participants received detailed written information about the project, and the modified JLA-process. They then provided their written informed consent, acknowledging that their participation was voluntary, and that they could withdraw from the project at any time.

### Consent for publication

Not applicable.

### Availability of data and materials

The data sets generated and analysed during the current study are not publicly available due to potential privacy concerns but are available from the corresponding author upon reasonable request.

### Author Contributions

AMNB had overall responsibility for the study and together with the rest of the project group, AMNB led participant recruitment and conducted all five stages of the modified JLA PSP process, including data collection, workshops preparation and facilitation. AMNB, GJ, SH, IS and DK contributed to the study’s conception and design, analysis, interpretation and critical revision of the manuscript. AMNB was the main author of the manuscript, with substantial editing and discussion of the manuscript by all authors. All authors read and approved the final version of the manuscript and agree to be accountable for all aspects of the work, ensuring that questions related to the accuracy or integrity of any part are appropriately investigated and resolved.

## Acknowledgements

The project group would like to express their gratitude to the participants in the steering group and the expert panel who contributed their knowledge and experience to aid in the process of developing top ten list of research topics. We also wish to thank the hospital ward leaders and the Department of Anaesthesia, who facilitated the recruitment process.

## Supporting information

**S1 Fig.1. A modified James Lind Alliance process**. (DOCX)

**S2 Text. Setting. REPRISE checklist for the priority setting process**. (DOCX)

**S3 Text. Setting. GRIPP-SF reporting checklist for documenting user involvement**. (DOCX)

**S4 Text. Setting. COREQ checklist in prepering the manuscript**. (PDF file)

